# Spot Urine Protein to Creatinine Ratio in Patients with Urinary Tract Infection

**DOI:** 10.1101/2024.12.29.24319746

**Authors:** Mukesh Chauhan, SV Suresh Babu

**Affiliations:** Padmashree Institute of Medical Laboratory Technology, Rajiv Gandhi University of Health Sciences, Bangalor, India; sri Siddhartha Medical College,Affiliated to sri Siddhartha Academy of Higher education,tumkur

**Keywords:** proteinuria, protein to creatinine ratio (P/C ratio), urine microprotein (UmiP), urinary tract infection (UTI)

## Abstract

**Introduction:** Untreated urinary tract infections (UTIs) can lead to complications, including renal deterioration due to upper urinary tract involvement. Proteinuria, characterized by excessive protein in the urine, is often indicative of kidney damage. The protein-to-creatinine ratio (P/C ratio) test is a convenient and reliable method for assessing proteinuria. This study aimed to evaluate the urine protein-to-creatinine ratio (UPCR) in UTI patients and its association with renal impairment.

**Materials and Methods:** Eighty patients with confirmed UTI and suspected proteinuria were recruited. Urine screening included pyuria (white blood cell presence) as an initial indicator of UTI, followed by microscopic examination of centrifuged urinary sediments. The urine supernatant was analyzed for protein using the urine strip method.

**Results:** After applying exclusion criteria, forty-six patients (n=46) were included in the statistical analysis. Of these, 26% had normal proteinuria (<15 mg/mM Cr), 35% had moderate proteinuria (15-50 mg/mM Cr), and 39% exhibited severe proteinuria (>50 mg/mM Cr). Patients were categorized into three stages (I, II, and III) with mean creatinine excretion values of 33.9 +/-13.9 mg/dL, 31.2 +/-17.2 mg/dL, and 29 +/-13.6 mg/dL, respectively, all significantly below the reference interval (168 +/-132 mg/dL).

**Conclusion:** Increased urinary protein excretion correlates with heightened risk of renal complications, a leading factor in mortality. Urinary protein excretion was markedly elevated in Stage III patients. The P/C ratio proved to be a more accurate diagnostic marker within the urine profile, highlighting proteinuria in UTI patients as a potential risk factor for renal impairment.

## Introduction

Urinary tract infections (UTIs) are among the most common bacterial infections, affecting approximately 150 million individuals globally each year [1]. These infections, which can involve any part of the urinary system, affect about 40% of women and 12% of men at least once in their lifetime, with around 40% of affected women experiencing recurrent UTIs [2]. If left untreated, UTIs can progress to the upper urinary tract, potentially leading to severe complications, such as sepsis, septic shock, and acute renal damage. Common symptoms include frequent, intense urges to urinate, as well as painful or burning sensations during urination [3, 4]. In severe cases, UTIs may lead to chronic kidney infections, which can cause irreversible kidney damage and even become life-threatening if bacteria enter the bloodstream (septicemia) [5].Proteinuria, the abnormal presence of protein in urine, is an important indicator of kidney damage and is often associated with UTIs [6]. Normally, albumin filtration occurs in the glomerulus, with subsequent reabsorption in the tubules. Dysfunction in either the glomerulus or proximal tubules disrupts this process, leading to increased albumin excretion, known as albuminuria [7, 8]. Proteinuria can be detected through various methods, including reagent strip tests in clinical settings and laboratory immunoassays for specific proteins such as albumin [9]. Screening for proteinuria is crucial for early detection of proteinuric kidney diseases.

In addition to being a marker for kidney disease, proteinuria serves as a prognostic indicator for progressive renal impairment and cardiovascular disease [10, 11]. For individuals with diabetic nephropathy, primary glomerulonephritis, or chronic kidney disease (CKD), urinalysis to assess proteinuria is a valuable tool in identifying those at increased risk for renal progression. Evaluating proteinuria is also recommended for hypertensive patients, as well as for early detection of diabetic nephropathy in diabetic patients [12, 13]. Even among non-diabetics, microalbuminuria functions as a cardiovascular risk marker [14]. Some studies suggest that UTIs are frequently associated with proteinuria; in fact, about three-quarters of confirmed UTI cases show positive reagent-strip tests for protein [15, 16]. Routine urine protein testing, therefore, not only aids in assessing kidney function but also provides insights into other conditions that result in proteinuria [17].

The protein-to-creatinine (P/C) ratio test, which can be conducted on a single-void (“spot”) specimen, has been shown to correlate closely with 24-hour proteinuria, as established by Ginsberg et al. [18]. This method has since been validated in numerous studies [19], and the Kidney Disease Outcome Quality Initiative (K-DOQI) recommends it as a reliable alternative to 24-hour proteinuria measurement [20, 21]. The P/C ratio is widely considered the most convenient and reliable method for detecting proteinuria. This study aims to evaluate the urinary protein-to-creatinine ratio (UPCR) in patients with UTI and assess its association with kidney damage.

## Materials and Methods

### Study design

This open-blinded, cross-sectional study was conducted in the Department of Internal Medicine at Padmashree Diagnostics, Vijayanagar, Bangalore, India. The study protocol was approved by the Institutional Human Ethics Committee (RGCB IHEC) of Rajiv Gandhi University of Health Sciences (registration no: DCGI - ECR/484/Inst/KL/2013, DHR-EC/NEW/INST/2020/477), ensuring compliance with ethical standards for research involving human participants.

### Patient recruitment and consent

Patients attending Padmashree Diagnostics for the diagnosis and treatment of urinary tract infections (UTIs) were recruited for this study through random sampling. Inclusion criteria required that participants be adults between the ages of 20 and 65, present with symptoms of UTI, and be drug-naïve for their infection. Exclusion criteria included those with a history of recurrent infections, multiple drug use, catheter-associated infections, pregnancy, and other conditions that could confound proteinuria results. All participants provided written informed consent prior to enrollment, and the study was reviewed and approved by the Institutional Review Board of Padmashree Institute of Clinical Research (PICR), Bangalore. The consent process included a detailed explanation of the study’s purpose, procedures, and potential risks [22, 23].

### Patient selection

A total of 80 patients with confirmed urinary tract infections (UTIs) and suspected proteinuria were recruited for this study through random sampling, conducted from August 2017 to June 2018 at the Department of Internal Medicine, Padmashree Diagnostics, Vijayanagar, Bangalore, India. Eligible participants were adults between 20 and 65 years, exhibiting symptoms consistent with UTIs and testing positive for proteinuria on initial urine strip tests. Patients were excluded if they had a history of recurrent infections, prior treatment for UTI, catheter-associated UTIs, pregnancy, or conditions like polycystic kidney disease, which could confound proteinuria assessment (see Table 1 for detailed inclusion and exclusion criteria. Of the initial 80 participants, 27 were excluded due to low creatinine levels (<20 mg/dL), and 7 were excluded due to the absence of proteinuria. This left a final sample of 46 patients for statistical evaluation, with data collection structured to minimize bias Of the initial 80 participants, 27 were excluded due to low creatinine levels (<20 mg/dL), and 7 were excluded due to the absence of proteinuria. This left a final sample of 46 patients for statistical evaluation, with data collection structured to minimize bias.

**Table 1.** Inclusion and exclusion criteria.

| **Inclusion criteria** | **Exclusion criteria** |
| --- | --- |
| Age: 20 – 65 years | Age: Below 20 years, above 65 years |
| Gender: Male and female | Patient with history of taking multiple drugs |
| Patients with clinical diagnosis of Urinary Tract Infection (Drug Naïve) | Patients with repeated infections & treated subjects |
| Symptomatic UTI | Catheter associated UTI |
| Urine strip positive for Protein | Pregnancy, polycystic kidney |
| Normal renal function | Inability to take oral medication |

### Specimen collection and storage

Urine specimens were collected under sterile conditions and sent to the microbiology department for urinary culture and sensitivity testing as part of the clinical procedures for this study. Additional specimens, initially collected for biochemical tests requested by attending physicians but no longer needed, were used for further analysis in this research. All samples were treated as abnormal/pathological and handled according to the study’s protocols.

Inclusion and exclusion criteria were strictly applied to minimize bias and ensure the quality of the specimens used in the analysis (see Table 1).

### Specimen assay

Urine samples were centrifuged at 1800 ×g for 15 minutes to separate cellular components. The resulting cell-free supernatant was aliquoted, labeled, and stored at -20°C until further analysis.

Screening for urinary white blood cells (pyuria), a primary diagnostic test for UTI, was conducted by microscopic examination of centrifuged urinary sediments. Approximately 10 mL of each urine sample was centrifuged at 2000 rpm for 5 minutes, and the sediment was then examined microscopically under both low- and high-power objective lenses for the presence of pyuria. Following this examination, the sediment was subjected to culture and sensitivity tests to determine bacterial presence and antibiotic susceptibility.The urine supernatant was then screened for protein using the urine strip method. Quantitative estimations of urinary protein and creatinine were performed by sulfosalicylic acid (SSA) precipitation and the spectrophotometric Jaffe reaction, respectively [13].

### PCR calculation

The protein-to-creatinine ratio (P/C ratio) is a standardized method to assess proteinuria in patients. It represents the amount of urinary protein relative to urinary creatinine, generally expressed as milligrams of protein per millimoles of creatinine. This study employed the following formula to calculate the P/C ratio:

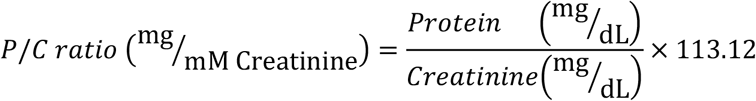

where:

- **Protein (mg/dL)** is the concentration of protein in the urine sample.
- **Creatinine (mg/dL)** is the urinary creatinine concentration.
- **113** is the conversion factor, used to express the P/C ratio in units of mg/mM, providing consistency with clinical standards of a spot urine sample for the P/C ratio was preferred over the 24-hour urine collection due to its ease, minimal patient burden, and the elimination of errors associated with incomplete collection or timing inaccuracies . This ratioeliable indicator for assessing kidney function in patients with urinary tract infections, aiding in the early detection of renal impairment.

## Results

### 1. Participant Demographics and Characteristics

Of the initial 80 participants, 27 were excluded due to low urinary creatinine levels (<20 mg/dL), and 7 others were excluded due to the absence of proteinuria. This left a final sample size of 46 participants, with a mean age of 38.11 ± 7.3 years. Of these, 67% were female (n=31) and 33% were male (n=15).

### 2. Classification of Proteinuria

Based on the KDIGO (2012) guidelines, participants were classified into three stages of proteinuria:

- **Stage I (Normal/Mild)**: Proteinuria <15 mg/mM creatinine (n=12)
- **Stage II (Moderate)**: Proteinuria 15-50 mg/mM creatinine (n=16)
- **Stage III (Severe)**: Proteinuria >50 mg/mM creatinine (n=18)

Among the 46 participants, 26% (n=12) were classified with normal proteinuria levels (<15 mg/mM creatinine), 35% (n=16) exhibited moderate proteinuria (15–50 mg/mM creatinine), and 39% (n=18) had severe proteinuria, with levels exceeding 50 mg/mM creatinine (Figure 5.3).

This categorization provided a structured approach to assessing renal impairment severity associated with urinary tract infections.

**Fig 5.3.**
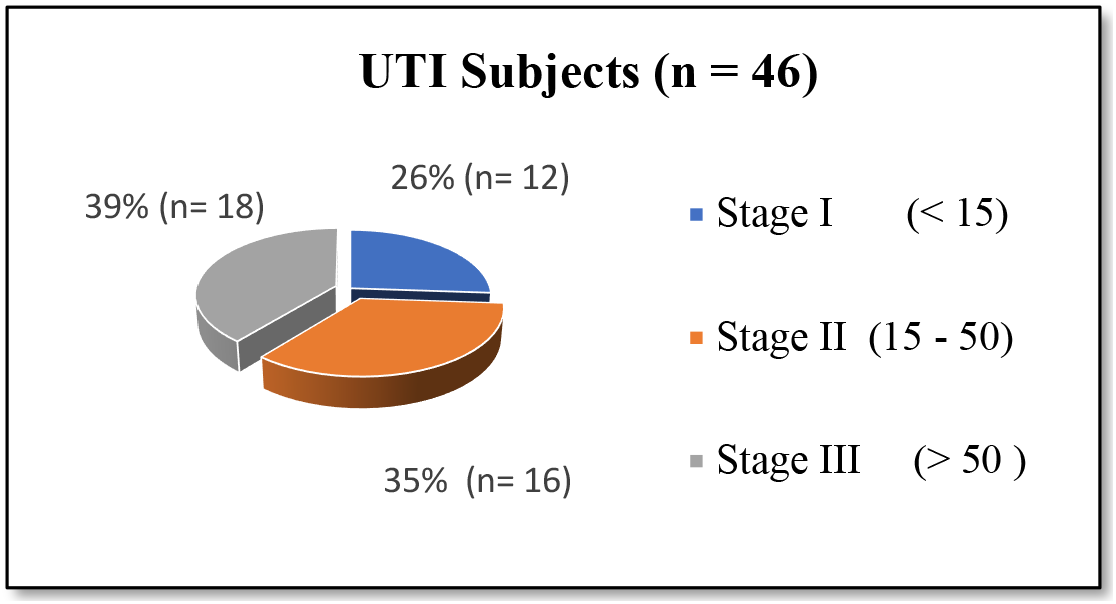
Diagramatic presentation of UTI subjects categorized in to 3 stages

**Table 2.** details the distribution of urinary protein, creatinine excretion, and P/C ratio for each stage:

| Stage | Urinary Protein (mg/dL) | Urinary Creatinine (mg/dL) | P/C Ratio (mg/mM Creatinine) |
| --- | --- | --- | --- |
| I (Normal/Mild) | $2.6 \pm 1.7$ | $33.9 \pm 13.9$ | $9.0 \pm 3.6$ |
| II (Moderate) | $8.0 \pm 4.6$ | $31.2 \pm 17.2$ | $30.4 \pm 8.0$ |
| III (Severe) | $48.5 \pm 46.0$ | $29.0 \pm 13.6$ | $180.6 \pm 142.9$ |

### 3. Urinary Creatinine Excretion

Urinary creatinine excretion was significantly reduced across all proteinuria stages compared to the reference interval (168 ± 132 mg/dL). Participants in Stage I, II, and III exhibited mean creatinine excretion values of 33.9 ± 13.9 mg/dL, 31.2 ± 17.2 mg/dL, and 29.0 ± 13.6 mg/dL, respectively (Figure 5.4). These values indicate a statistically significant [24, 25] decrease (p < 0.001) in urinary creatinine among patients with suspected urinary tract infections, suggesting an impact on renal clearance.This progressive decline in creatinine excretion highlights a potential disturbance in renal clearance mechanisms associated with UTIs, affecting overall kidney function. Urinary creatinine, a widely accepted diagnostic and prognostic marker, serves to normalize biomarker excretion for a more precise assessment of renal impairment. In this study, the observed reduction in urinary creatinine underscores the diagnostic utility of creatinine normalization in detecting renal dysfunction in UTI patients [26, 27].

**Figure 5.4.**
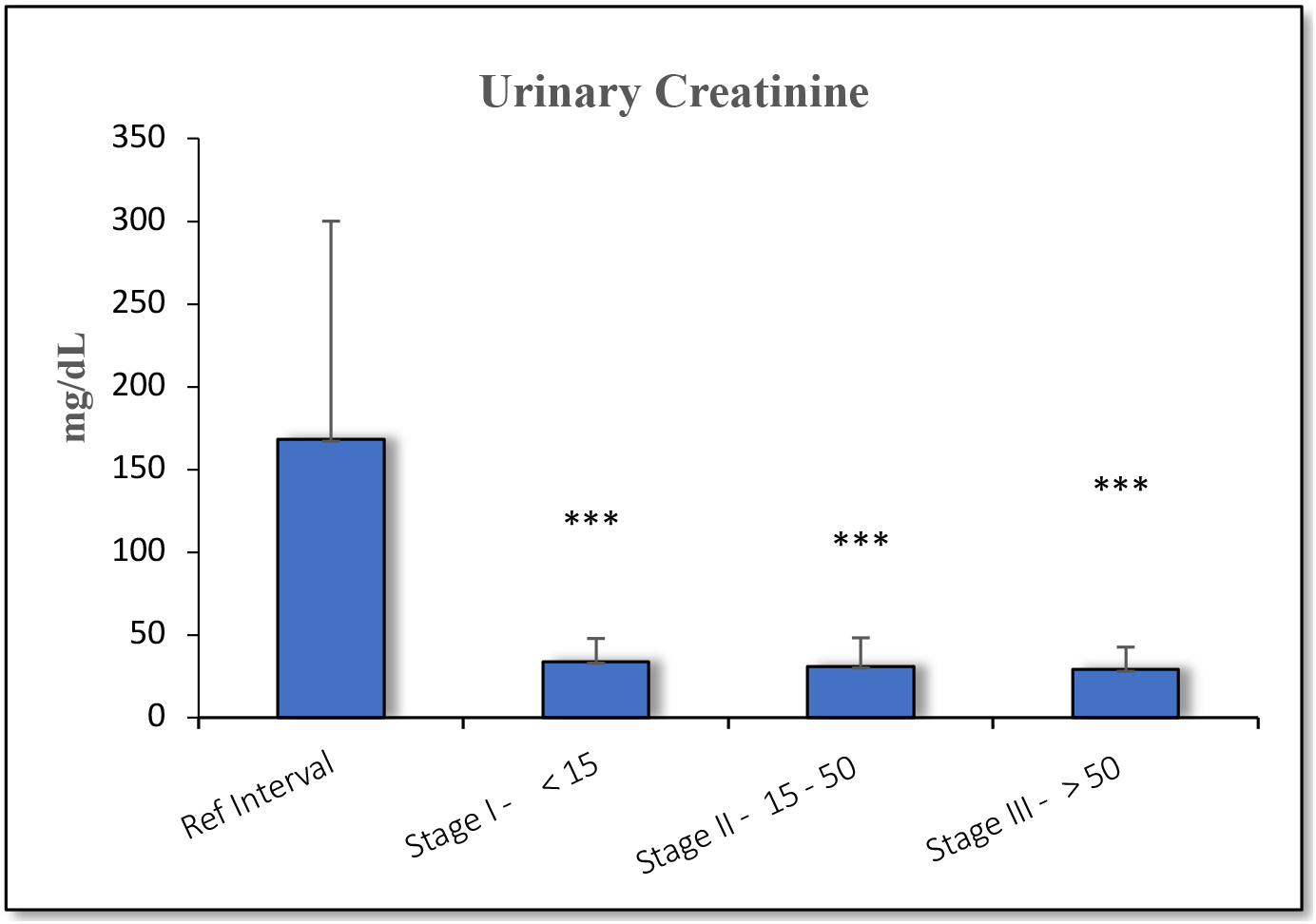
Histogram representing the urinary creatinine values among the categorized UTI subjects. Values expressed as mean ± SD. Student’s ‘t’ test: *: p <0.05; **: p<0.01; ***: p< 0.001.

### 4. Urinary Microprotein Levels

Urinary microprotein has been identified as a biomarker for the progression of urinary tract infections (UTIs), especially in cases with comorbidities such as insulin-resistant diabetes, renal infections, and hypertension. In the current study, Stage III participants exhibited a significantly elevated urinary microprotein excretion level (48.6 ± 46 mg/dL) compared to the normal reference interval (9.0 ± 2.6 mg/dL), as illustrated in Figure 5.5. This increase was statistically significant (p < 0.001) [28, 29].

**Figure 5.5.**
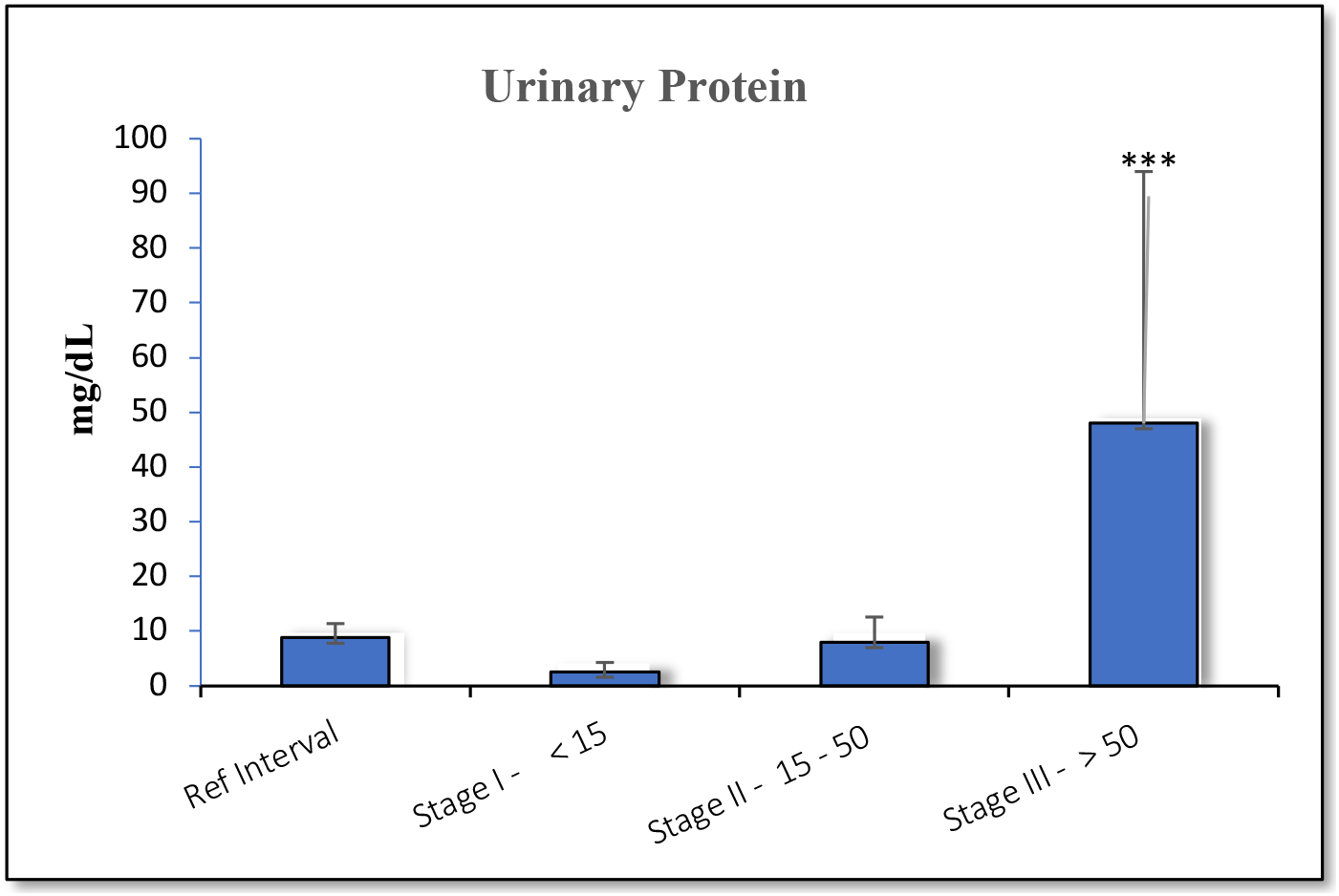
Histogram representing the urinary protein among the categorized UTI subjects. Values expressed as mean ± SD. Student’s ‘t’ test: *: p <0.05; **: p<0.01; ***: p< 0.001.

The observed elevation in urinary microprotein suggests that UTIs may adversely affect renal function, with urinary protein excretion potentially serving as an indicator of disease severity. Analysis of Figures 5.4 and 5.5 revealed that protein excretion levels increased steadily across stages, contrasting with the decrease in creatinine excretion. This pattern may reflect a worsening renal prognosis, as increased urinary protein excretion is often associated with deteriorating kidney health.

The findings support the utility of spot urinary protein measurements for understanding the pathophysiology of UTI-related disease progression. These results indicate that urinary protein excretion may serve as a valuable prognostic marker, warranting further longitudinal studies to evaluate its clinical significance and to assess therapeutic outcomes over time.

### 5. Protein-to-Creatinine Ratio (P/C Ratio)

The protein-to-creatinine (P/C) ratio emerged as a sensitive indicator of renal dysfunction in UTI patients. In particular, Stage III participants demonstrated the highest mean P/C ratio (180.6 ± 142.9 mg/mM creatinine), which was significantly elevated compared to the lower stages (Figure 5.6). This increase in the P/C ratio across stages was statistically significant, affirming its relevance as a diagnostic tool for assessing kidney impairment severity.

**Fig 5.6.**
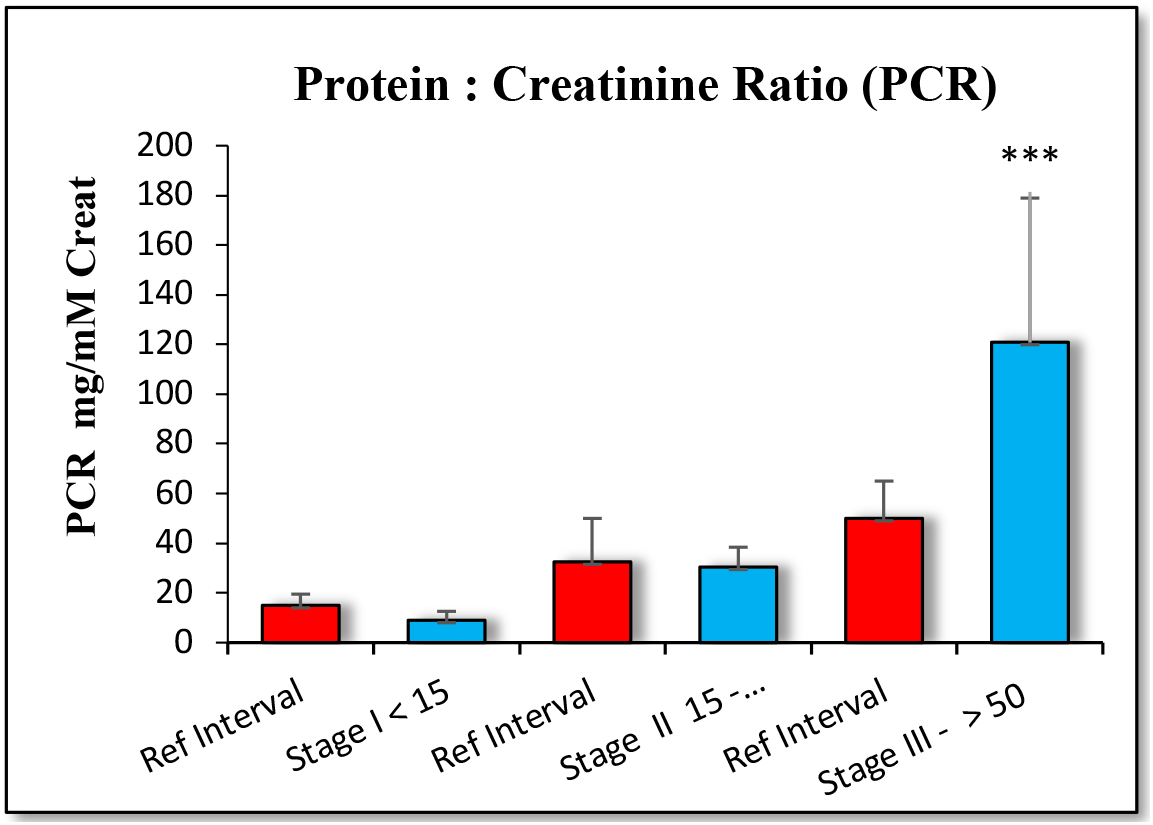
Histogram represents the PCR among the categorized UTI subjects. Values expressed as Mean ± SD. Student’s‘t’ test: *: *p* <0.05; **: *p*<0.01: ***: *p*< 0.001

Among urinary biochemical markers, derived parameters such as the P/C ratio and Albumin-to-Creatinine Ratio (ACR) offer improved diagnostic utility. These ratios provide a more reliable indication of underlying renal pathology, especially when individual values of urinary protein or creatinine are within reference ranges but the combined ratio reveals pathology. The P/C ratio thus plays a crucial role in enhancing diagnostic accuracy, offering a robust index of disease severity in cases where standard markers may be ambiguous.These findings support the use of the P/C ratio as a clinically valuable parameter for identifying and monitoring kidney impairment in UTI patients, underscoring its significance in evaluating disease progression and renal function.

## Discussion

Biochemical analysis of urinary protein and urinary creatinine is a critical indicator for diagnosing renal impairment and assessing urinary tract infections (UTIs) [30]. Early screening for UTI is essential to understanding the disease's pathophysiology, as delayed diagnosis may lead to irreversible damage [31]. In UTI patients, a comprehensive evaluation—including urine routine tests, microscopic examination, and measurements of nitrite, protein, leukocyte esterase, albumin, and creatinine—provides insights into disease progression and treatment response. Additionally, urinary creatinine levels correlate with sex, age, and BMI, reflecting its role as a proxy for muscle mass in patients with chronic kidney disease (CKD) [32, 33].

Our study results indicated a statistically significant reduction in urinary creatinine excretion among UTI patients, aligning with findings by Tynkevich et al., who reported decreased 24-hour creatinine excretion in early CKD stages [34]. Although there were significant differences in creatinine levels between groups and the normal reference, no significant differences were observed among the UTI groups themselves (Figure 5.4), underscoring that urine creatinine alone may not adequately differentiate between disease stages. On the other hand, urinary microprotein excretion was significantly elevated in Stage III (48.6 ± 46 mg/dL) compared to the normal reference interval (9.0 ± 2.6 mg/dL), suggesting impaired glomerular filtration and possible reabsorption deficiencies (Figure 5.5). Despite the lack of endorsement for proteinuria as a surrogate endpoint in CKD clinical trials by the FDA and European Medical Agency, accumulating evidence has spurred discussions among nephrologists and FDA representatives regarding its potential utility as a prognostic marker [35].

Traditionally, 24-hour creatinine testing is used due to diurnal variations in creatinine excretion [36]. However, 24-hour collection is logistically challenging, often yielding errors from inconsistent timing, bacterial contamination, incomplete collections, and insufficient bladder emptying. These issues, along with the cost and inconvenience of hospitalization for follow-up, have led to the consideration of alternatives [36]. The spot urine protein-to-creatinine ratio (P/C ratio) offers a reliable and convenient alternative for proteinuria estimation, with a strong correlation to 24-hour protein excretion, as confirmed by Rodriguez-Thompson et al. [37].

Our findings support the use of the P/C ratio as an effective diagnostic marker, as it reflects UTI impact on both upper and lower urinary tracts. Among urinary biomarkers, derived ratios such as P/C and albumin-to-creatinine ratio (ACR) demonstrate superior diagnostic value [38]. The present study classified UTIs into three stages based on P/C ratios: Stage I (<15 mg/mM creatinine), Stage II (15–50 mg/mM creatinine), and Stage III (>50 mg/mM creatinine). This classification highlights the diagnostic utility of the P/C ratio, especially in patients with normal serum creatinine and proteinuria levels but impaired creatinine clearance (Figure 5.4). Screening based on P/C ratio may facilitate earlier identification of high-risk individuals, thus enhancing management strategies for both lower and upper urinary tract health.These findings underscore the necessity for sensitive diagnostic guidelines, such as those in the CKD 2012 KDIGO recommendations. Future studies should explore the prognostic significance of urinary protein excretion by conducting longitudinal assessments to monitor therapeutic outcomes in UTI patients.

## Conclusion

This study demonstrated a significant association between proteinuria and urinary tract infections (UTIs). Urinary creatinine levels were notably reduced across all stages of disease severity compared to the normal reference interval, indicating potential impairment in renal function due to UTIs. This reduction suggests that monitoring urinary creatinine can serve as a benchmark in pharmacotherapy, aiding in the assessment of glomerular filtration and the accuracy of drug clearance monitoring.

Additionally, urinary protein excretion was significantly elevated in the Stage III disease group, reflecting severe impairment in renal filtration and reabsorption processes. The measurement of urinary creatinine and protein excretion together enhanced the diagnostic sensitivity for detecting proteinuria, offering a clearer understanding of disease severity. Importantly, the protein-to-creatinine (P/C) ratio was identified as a more reliable diagnostic marker within the urinary profile, providing valuable insights into the progression of UTI-associated renal impairment.

These findings underscore the importance of incorporating the P/C ratio in routine diagnostic evaluations, particularly in patients at higher risk of renal complications from UTIs, as it offers a robust indicator of disease progression and therapeutic response.

## Data Availability

All data produced in the present study are available upon reasonable request to the authors

## Appendix

**Figure 5.6.**
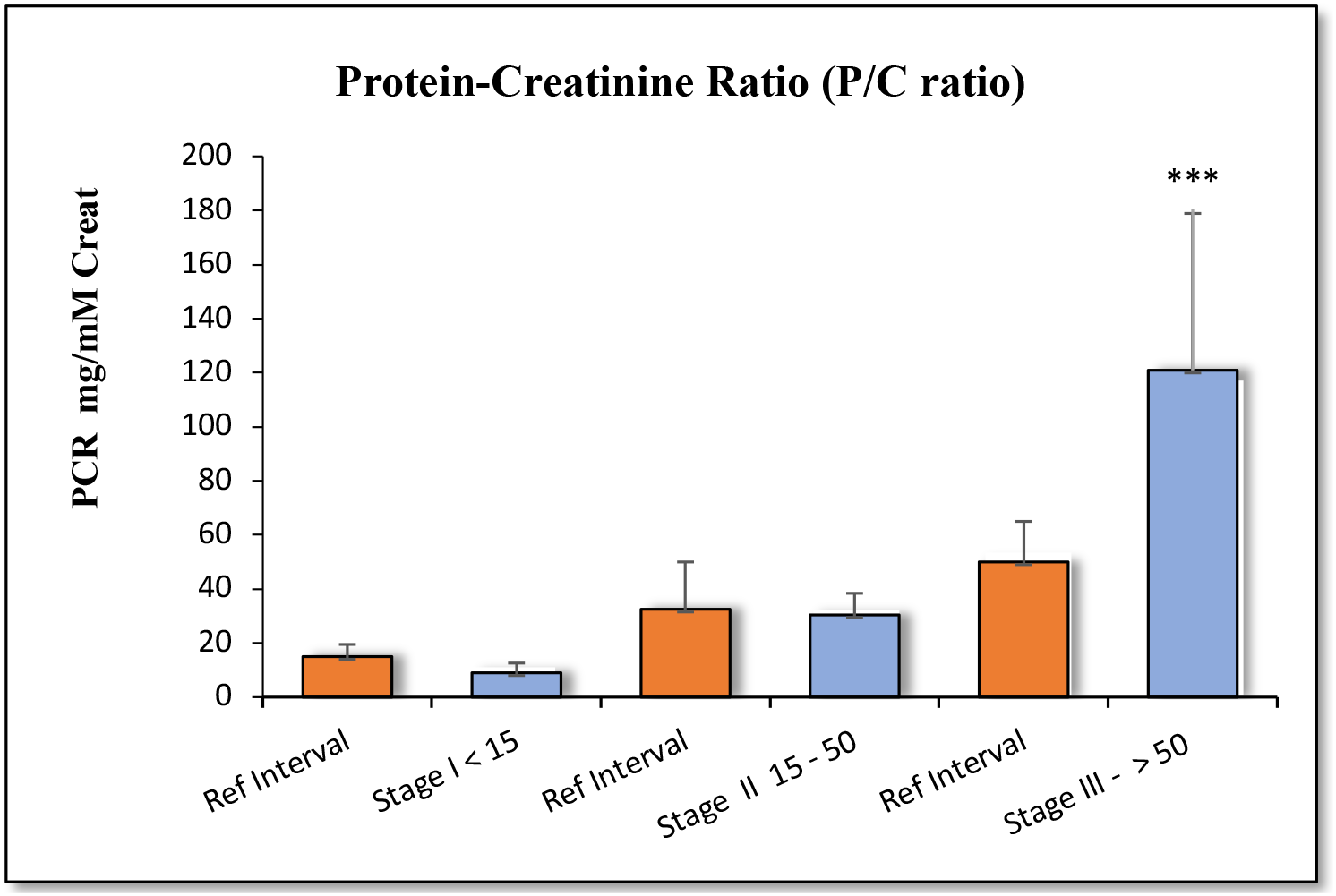
Histogram representing the P/C ratio among categorized UTI subjects. Values expressed as mean ± SD. Students ’t’ test: *: p <0.05; **: p<0.01; ***: p< 0.001.

